# Exploring the knowledge and acceptance of reputed Authorship Criteria: A Pilot Study among medical researchers in India

**DOI:** 10.1101/2022.06.10.22276195

**Authors:** Bhavik Bansal

## Abstract

**Objectives:** To determine knowledge and acceptance of authorship criteria among residents, PhD scholars and faculty involved in medical research in India.

**Design:** A cross sectional survey was performed via Google forms (a web based platform).

**Results:** A total of 117 participants responded to the survey, of whom 66 (56%) were faculty/professors, 23 (20%) residents and 28 (24%) PhD scholars. 33% respondents had faced conflicts with their guide, 58% respondents have offered honorary authorship sometime in their careers. Only half of the respondents were aware of the ICMJE guidelines for authorship.

**Conclusions:** Gift Authorship and ‘pressure to publish’ are largely prevalent in bio medical research in India. Journals requiring author-contribution declarations, overlooking the number of publications as the sole source of offering academic promotions among others are possible solutions to curb this problem.

## INTRODUCTION

Research presents as the ultimate tool to add to the existing knowledge on a topic. A research report is a free communication by a scientist or a group of scientists informing their peers about a set of novel findings that either provide answers to puzzling problems or raise issues that are of academic or practical interest.

However, recently the idea of publishing papers has sadly come to be driven by the increased chances of getting hired and getting academic promotions rather than the will to add to science itself. In medical research these days, the line between a credit-worthy contribution and the ‘honour’ of being a co-author is starting to fade. Institutional medical research in developing countries like India is seeing a rise in the tendency among authors to award authorship as a measure of one’s respect towards a colleague (here, called a ‘Guest Author’). The work of such a guest author is referred to as ‘Honorary Authorship’.

As ‘authorship’ becomes a more sought after tag, published papers are seeing an increase in the list of authors and hence the prevalence of “honorary authorship” in recent years. (1) The act of offering gift authorship is even more common among junior authors (2), who are commonly seen appending the name of the head of their labs/departments as co-authors on their papers for getting the required funds for their project and/or getting access to research subjects.

In addition to awarding authorship to a renowned name in the field to garner credibility, if the real contributors behind the work are hidden which may or may not be to prevent an impending conflict of interest, the scientific community is left with a lack of information behind this ‘Ghost Author’s’ affiliations and ulterior motives. This practice is often called ‘Omitted authorship’. (3,4) The debate over order of authors as well as responsibilities too has drawn attention of many.

In this paper we have focused on the prevalence of malpractices surrounding authorship, tried to dig at reasons for the same and evaluated factors that could possibly lead to it, including awareness of the reputed ICMJE criteria.

### ICMJE guidelines

The International Committee of Medical Journal Editors (ICMJE) defines an author as an individual who has made substantial intellectual contributions to a scientific investigation and fulfills the following criteria :

1. *Substantial contributions to the conception or design of the work; or the acquisition, analysis, or interpretation of data for the work (Scholarship); AND*
1. *Drafting the work or revising it critically for important intellectual content (Authorship); AND*
2. *Final approval of the version to be published (Approval); AND*
3. *Agreement to be accountable for all aspects of the work in ensuring that questions related to the accuracy or integrity of any part of the work are appropriately investigated and resolved (Agreement)*.

*In addition to being accountable for the parts of the work he or she has done, an author should be able to identify which co-authors are responsible for specific other parts of the work and have confidence in the integrity of the contributions of their co-authors*.

As per the guidelines, those who do not meet the aforementioned criteria must be acknowledged in a separate section. (5)

## REVIEW OF LITERATURE

Authorship fraud is the most prevalent of research frauds, among others such as data fabrication, plagiarism, data falsification, authorship fraud, publication fraud, and grant fraud. (6) A prevalence of honorary authorship ranging from 4% - 74% across various article types and high impact journals has been reported in literature. (7–10)

Gift authorship is often justified against the background of professional/non professional relationships that may have existed in the past or happen to exist in the present. It’s not uncommon to find seniors or colleagues from the same department getting listed as co authors on a paper. (11) Consequently, negative collaborative experiences are less commonly reported when the collaborators are from different universities as compared to those from the same university. (12)

The first author may find it harmless to offer the authorship as they shall still bear the tag of being the first author anyway and that ‘invoking professionalism’ would make them look outrightly self righteous when all it would take is to add another name in the byline. (13) Authorship gifting behaviors may even arise out of career building reasons which continually impose a pressure to publish, colloquially called the ‘Publish or Perish’ principle. (14) Order of authorship could only seldom predict the nature of contribution in papers published in leading biomedical journals. (15)

The fraction of articles, across various article types and study settings, with all co-authors fulfilling the ICMJE guidelines has been found to range between 64% - 68% in literature. (16–19). Self- reported awareness of the ICMJE guidelines was found to vary from 81.4% to 40%. (11,18,20–22) Even among the significantly small proportion of people who are able to specify all the three criteria, hardly few know that all criteria have to be met. (22,23) Among those who are not fully aware of the ICMJE guidelines, Statement 1 (“*Substantial contributions to the conception or design of the work; or the acquisition, analysis, or interpretation of data for the work”*)was agreed to be a part of the ICMJE criteria and Statement 3 (“the final approval of the manuscript version to be submitted for publication is required for authorship”) is least thought of as being a part of the criteria. (24)

## AIMS AND OBJECTIVES

### Primary Objective

1. To determine the prevalence of honorary authorship self reported by medical researchers in India, separately for faculty, PhD scholars and residents.
2. To determine the prevalence of conflicts among co-authors and the principal investigator regarding authorship order and/or inclusion.
3. To determine the awareness about reputed guidelines for author selection, like the ICMJE guidelines.

### Secondary Objective

1. To gauge the broader reasons people believe it is justified to offer honorary authorship.
2. To determine if a correlation exists between education abroad with knowledge and acceptance towards good authorship selection practices.
3. To determine if a correlation exists between formal research ethics training with knowledge and acceptance towards good authorship selection practices.

## MATERIALS AND METHODS

A web based platform called Google Forms was used to gather data from the questionnaire (attached at the end of the report). The data was collected over July-August 2021. The link to the form was disseminated through relevant groups, forums and mailing lists. The participants filled the form in an anonymised fashion, and therefore follow-up could not be performed. The sampling was done through a convenience based - non probability method.

Section 1 of the questionnaire is related to demographic details of the respondent, which are entirely anonymised, but identifies the respondent based on his/her experience in research. Section 2 builds more on prior experiences, including exposure to authorship guidelines, formal training in research ethics and part of education being abroad (defined as training lasting more than 6 months amenable to certification).

Section 3 focuses on the knowledge and views among respondents regarding currently accepted authorship guidelines. It comprises a 10-questions likert type questionnaire asking for opinions on statements prevalent as myths regarding authorship guidelines. Each respondent is given a score between +10 and -10 (it has been called the Section 3 Score in the manuscript) based on their (dis)agreement to a particular question. These scores are then analysed in context of their demographics and prior experiences.

## RESULTS

### Sample Characteristics

The study included 117 participants of whom 66 (56%) were faculty/professors, 23 (20%) residents and 28 (24%) PhD scholars. Professors who responded to the questionnaire had a median experience of 14.5 years, and a median of 41.5 publications till date with 26% (n=17) having received formal training abroad, while residents and PhD scholars had more than a median experience of 5 years, and more than a median of 4 publications. (Table 1.1)

**Table 1.1:** Demographic characteristics of sub-groups; Table 1.2 : Prior experiences among participants; Table 1.3 : Awareness about the ICMJE Guidelines among participants; Table 1.4 : Source of first exposure to authorship guidelines among participants; Table 1.5 : Role in current projects of the participants

| Group | Faculty/Professor (n = 66) | Resident (n = 23) | PhD Scholar (n = 28) | Total (n = 117) |
| --- | --- | --- | --- | --- |
| Sample characteristics |  |  |  |  |
| Frequency (% Total) | 66 (56%) | 23 (20%) | 28 (24%) | 117 (100%) |
| Experience - Median (IQR) | 14.5 (10.0 - 23.5) | 5.0 (1.0 - 8.0) | 5.5 (3.25-7.75) | 10.0 (5.0 - 20.0) |
| Number of Publications - Median (IQR) | 41.5 (18.5 - 150.0) | 4.0 (3.0 - 8.0) | 4.5 (1.25 - 16.25) | 17.0 (4.0 - 50.0) |
| **Prior Experiences** |  |  |  |  |
| Formal training abroad | 17 (26%) | 0 (0%) | 2 (7%) | 19 (16%) |
| Conflict with guide regarding authorship | 21 (32%) | 9 (39%) | 9 (32%) | 39 (33%) |
| Offered honorary authorship | 35 (53%) | 14 (61%) | 19 (68%) | 68 (58%) |
| Formal Course on Research Ethics | 42 (63%) | 9 (39%) | 11 (39%) | 62 (53%) |
| **Awareness about the ICMJE Guidelines** |  |  |  |  |
| Yes, I am aware | 45 (68%) | 9 (40%) | 5 (18%) | 59 (50%) |
| Not aware | 9 (14%) | 7 (30%) | 12 (43%) | 28 (24%) |
| I am aware they exist. (I have not read them) | 12 (18%) | 7 (30%) | 11 (39%) | 30 (26%) |
| **Source of first exposure to authorship guidelines** |  |  |  |  |
| Mentor/Guide | 25 (38%) | 8 (35%) | 10 (36%) | 43 (37%) |
| Colleague | 4 (6%) | 0 (0%) | 4 (14%) | 8 (7%) |
| Internet | 11 (17%) | 5 (22%) | 4 (14%) | 20 (17%) |
| Formal Training | 19 (29%) | 1 (4%) | 1 (4%) | 21 (18%) |
| I don't have any prior exposure. | 7 (10%) | 9 (39%) | 9 (32%) | 25 (21%) |
| **Role in current Project** |  |  |  |  |
| Data Analysis | 29 (44%) | 18 (78%) | 27 (96%) | 74 (63%) |
| Data Collection | 25 (38%) | 16 (70%) | 21 (75%) | 62 (53%) |
| Experimental Design/<br>Innovation | 41 (62%) | 9 (39%) | 21 (75%) | 71 (61%) |
| Manuscript Writing/<br>Editing | 49 (74%) | 18 (78%) | 26 (93%) | 93 (79%) |
| Clinical Intervention | 27 (41%) | 9 (39%) | 7 (25%) | 43 (37%) |
| Wet Lab Work | 10 (15%) | 5 (22%) | 19 (68%) | 34 (29%) |

### Authorship

33% respondents had faced conflicts with their guide regarding inclusion or order of co-authorship. 58% responders (35 Professors, 14 residents and 19 PhDs) admitted to having offered honorary authorship (Table 1.2) out of several reasons viz. seeking a proofreading from the person concerned as they are more experienced (n=67), as a token of respect/legacy (n=54), it kept the team motivated and helped maintain good relations (n= 48), the probability of publishing increased (n= 38), they bring funding for the research and more opportunities to the table (n=21), felt a pressure to publish (n= 15). (Table 2)

**Table 2:** Reasons for Honorary Authorship as reported by study participants.

| **Reason for Honorary Authorship (n = 117)** |  |
| --- | --- |
| There is always a pressure to publish. | 15 (13%) |
| They bring in money/opportunities. | 21 (18%) |
| They help in proofreading because they have immense experience. | 67 (57%) |
| Out of respect and/or legacy to do so. | 54 (46%) |
| That way the probability of publishing increases. | 38 (32%) |
| Motivates team/maintain relations. | 48 (41%) |

On being questioned on their current knowledge of authorship criteria and ICMJE guidelines, only 50% were fully aware of the ICMJE guidelines while the rest were either not aware they existed (24%) or had not read them (26%). Only 53% of the responders had had formal training on research ethics. (Table 1.3) Mentors/ guides were the source of information regarding authorship guidelines and research ethics for 43 (37%) of the respondents. (Table 1.4)

### Current project

Of those currently working on a project, most respondents 73% (n= 93) reported to be involved in manuscript writing/editing. Residents and PhD scholars were more likely to be involved in data collection and data analysis (>70%), faculty were more likely to be involved in experimental design/innovation (62%). PhD scholars were actively involved in almost all areas of research work-data analysis (96%), data collection (75%), experimental design/innovation (75%), Manuscript writing (93%) and Lab work (68%) (Table 1.5).

### Knowledge Score

The distribution of Section 3 scores was roughly a gaussian distribution centred at the mean of -0.07 with a standard deviation of 3.15. The score for sub-groups based on formal training abroad, formal training in research ethics and awareness about ICMJE guidelines showed a positive, yet not a significant change. (Table 3)

**Table 3:** Knowledge Scores among subgroups classified as one who responded differently to questions as in different rows.

| **Sub-Groups based on :** | **Yes - Mean (Z value)** | **No - Mean (Z Value)** |
| --- | --- | --- |
| **Formal training in research ethics** | + 0.72 (0.251) | - 1.08 (-0.321) |
| **Education abroad** | + 2.1 (0.689) | - 0.53 (-0.146) |
| **Aware about ICMJE Guidelines** | + 0.44 (0.162) | - 0.64 (-0.181) |

## DISCUSSION

This survey was aimed at determining the prevalence of honorary authorship among residents, faculty and PhD scholars in India and gauging their awareness on the ICMJE authorship criteria. Honorary authorship was self-reported by 58% of our respondents of which maximum were PhD scholars (68%) and least were the faculty (53%). More than 40% based their offer of honorary authorship on the ground that their paper gets a proofread from a more experienced person, out of respect and/or that it motivates the team and maintains relations. Nevertheless, these results are not shocking enough when compared with similar studies.

Literature highlights various other difficulties around the idea of authorship which include being forced to add an undeserving author, (20) and exclusion from authorship when it was apparently deserved. (7,23) The results of our study show that conflicts among co-authors and the principal investigator regarding authorship order and/or inclusion are prevalent (>30% in each subgroup). Absence of a set mechanism to decide the order of authorship could be a reason behind this. (20). The lack of awareness of the authorship criteria as a reason for these conflicts was debunked by Dhaliwal et. al. (21) In our study, maximum negative collaborative experiences were reported by residents (39%) which could possibly be explained by their relatively less powerful position in academia. (12).

Only 50% of our respondents self-reported to be aware of the ICMJE criteria while 26% knew they existed but had not read them or were not aware of the guidelines at all (24%). These results fall in line with the existing literature as detailed in the review of literature.

Source of first exposure often sets the bar for behaviours that are expected and rewarded in academic research and defines the concept of ‘unacceptable deviation from shared norms of conduct.’ In India, graduate students get exposed to academic research mostly via a mentor or a guide in some department of their university. However departments do not generally make an effort to make students able to recognise and deal with ethical issues. (25,26) Yet again, the question persists, how significant an impact the training would make, even if undertaken. In our study neither formal training abroad nor formal courses on research ethics were found to make a significant impact on the knowledge of the authorship guidelines. Similar results have been reported in previous studies. (25) This low level of awareness among medical researchers possibly poses a challenge on ethical practising of authorship and has been used to explain the much prevalent misconduct in the scientific community in recent times. (11)

However, there are studies showing that level of awareness of the authorship criteria has little influence on the practice of authorship, and that more latent issues like the highly competitive environment and the absence of a check-and-report system in most institutions demand redressal. (7,21) More so, there are those who consider the guidelines rigid with regards to the practical barriers and the restrictive nature of the three listed criteria in the same. This has brought in criticism for the ICMJE which in turn has been used to ‘justify’ the ‘scientific misconduct’ with many authors debunking them entirely. (23). Authors have even reported contributions that could not be matched by any ICMJE criterion. (27) This has sparked a debate in academia, with many authors coming forward to support the ICMJE and calling the criticisms unjustified. (28)

### Strengths and Limitations

Our results might not be generalised due to the small sample size (n= 117) of our study, restricted due to the timed nature of the project and compliance among participants being lower than expected. Subjective nature of the scoring done in Section 3 could serve a grey area for our study. A study like ours, in the context of medical researchers in India wasn’t found in literature, which makes it a pilot study, therefore a sample size couldn’t be calculated pre-emptively. The sampling was done through a convenience based system, with a possible skewed distribution towards researchers in tertiary centres in Northern India.

Most of the markers of knowledge regarding authorship guidelines were derived using subjective statements, although well-established in literature, the statements used were not part of a previously validated questionnaire. Hence, interpretation of these results are subject to further validation of the study design, and implementation over a much larger sample size.

The questionnaire was administered among varied strata of medical researchers, while maintaining complete anonymity by ensuring that no question asked can trace the participant or their workplace/guide. This kind of anonymity encourages participants to be more open about their experiences regarding conflicts and disagreements with their superiors and colleagues.

This study therefore serves towards the initial efforts to bring attention towards the inconspicuous problem of authorship conflicts and publication pressure, encouraging further studies on these issues, particularly so in the context of medical researchers in India.

## CONCLUSION

Over the past couple years, research misconduct has gained substantial attention. Our study adds to it by having separate analyses for all the three subgroups involved (Residents, Faculty and PhDs scholars) and focusing on a sub-group less studies i.e of medical researchers in India. Differences were evident among the three groups. Different reasons could prevail, and need further investigation. It reinforces that gift authorship is largely prevalent in the scientific community.

Honorary authorship not only affects academic growth by invalidating records of indexing and citation tracking services but also releases certain crucial safeguarding checks on data reliability. (29) It puts a question mark on the accuracy and integrity of the new research papers being published. As this directly poses a threat to sound clinical decision making and policy formation, the problem needs prompt and a proper redressal mechanism. Various strategies have been discussed in literature to overcome this problem.

Supportive attention and prompt, detailed feedback may help circumvent many ethical problems. (26) Informal confidential channels such as Ombuds Office (18) and opportunities to discuss such issues openly will indirectly allow for corrective measures without compromising on any individual’s career. Getting a signed statement of justification for authorship/specifying their contributions in the paper and having a fixed-credit system could curb the prevalence of gift authorship, though some studies show non-significant effects of such systems already in place. (8)

Efforts should be made to reduce the ‘pressure to publish’ on medical researchers. Abolishing the concept of valuing a doctor’s or an institution’s quality in terms of its research output, and judging their research performance using a mere numerical value as h index is certainly needed. (30) Putting a cap on the number of publications in curriculum vitae could be another way towards curbing this pressure. (23) We predict that with the ongoing heated debate within the scientific community could eventually lead to a common solution that is agreeable to all.

## Data Availability

All data produced in the present study are available upon reasonable request to the authors.

## ACKNOWLEDGMENTS

We would like to thank Dr. Nalin Mehta (Professor, Department of Physiology, AIIMS Delhi) for mentoring in this ICMR-STS project. We would also like to thank Bhavya Kansal, who gave insightful comments to the manuscript drafting process.

## QUESTIONNAIRE/STUDY TOOL

***Available at*** : https://forms.gle/4FforS6mQcReYRMp8

## Notes

### Competing Interest Statement

The authors have declared no competing interest.

### Funding Statement

The study was done under the Short Term Studendship (STS) - 2019-20 program by Indian Council of Medical Research (ICMR).

### Author Declarations

Institute Ethics Committee for Post Graduate Research of All India Institute of Medical Sciences, New Delhi gave ethica, approval for this work.

